# “Each moon we come to weigh the pregnancy.” Self-efficacy in group antenatal care in Benin

**DOI:** 10.1101/2025.06.10.25329377

**Authors:** Julie Niemczura Sutton, Fifamè Aubierge Eudoxie Kpatinvoh, Esther Firmine Cadja Dodo, Erin Go, Courtney Emerson, Catherine Dentinger, Stephanie Suhowatsky, Julie Buekens, Katherine Wolf, Manzidatou Alao, Marie Adeyemi Idohou, Faustin Onikpo, Cyriaque D. Affoukou, Aurore Ogouyèmi-Hounto, Julie R. Gutman, Peter J. Winch

## Abstract

Group antenatal care (G-ANC) is a model that convenes women of similar gestational age for participatory education and antenatal care. A qualitative study was embedded in a trial assessing the impact of G-ANC on ANC retention and intermittent preventive treatment of malaria in pregnancy (IPTp) uptake to assess women’s experience of G-ANC, and ways participation could foster self-efficacy. Ten semi-structured focus group discussions were conducted with 129 women who attended G-ANC; deductive thematic codes were informed by Bandura’s four sources of efficacy expectations. Recently pregnant women’s experiences with individual ANC versus G-ANC were assessed via household surveys.

G-ANC participation proffered three sources of self-efficacy expectations: performance accomplishments, verbal persuasion, and vicarious experience. Among household survey respondents, 96% (134/140) of women who participated in G-ANC would prefer it over individual ANC for future pregnancies. While a higher proportion of G-ANC participants felt that the provider answered all their questions in a way they could understand, most women reported that not all their questions were answered.

G-ANC processes fostering self-efficacy to overcome barriers to ANC attendance may have facilitated women’s participation in G-ANC meetings as well as taking more doses of IPTp. Self-efficacy of pregnant women participating in G-ANC could be strengthened by providers addressing all participants’ questions in a more complete and understandable way, contributing to more effective verbal persuasion. Other parallel processes during G-ANC need to be maintained to provide multiple sources of self-efficacy for behaviors like emergency care-seeking, pregnancy management, pregnancy self-care, and facility birth.

## Introduction

Group antenatal care (G-ANC) is a service delivery model in which pregnant women with similar due dates are seen together in a group. This model was designed to promote adoption of key behaviors to increase the probability of healthy pregnancy outcomes (J. Sharma, O’Connor, & Rima Jolivet, 2018; Sheeder, Weber Yorga, & Kabir-Greher, 2012). Participatory education delivered by providers during G-ANC covers myriad health topics, helping women address barriers to successfully seek family planning, prevent or obtain treatment for malaria, take early action in response to danger signs during pregnancy, and deliver at a health facility. According to Grenier et al.’s theory of change, the structure and conduct of G-ANC meetings may influence maternal, newborn, and reproductive health behavioral outcomes by strengthening women’s health literacy and self-efficacy (Grenier et al., 2022). Recent studies indicate that G-ANC can increase knowledge, confidence, and empowerment (Oka, Madeni, & Horiuchi, 2022; Patil et al., 2017; Somji et al., 2022; Thapa et al., 2019); the likelihood of women having four or more antenatal care contacts (ANC4) (Grenier et al., 2019; McKinnon et al., 2020; Patil et al., 2017), uptake of interventions for malaria in pregnancy (Noguchi et al., 2020), and postpartum family planning use (Eluwa et al., 2018; Lori, Chuey, Munro-Kramer, Ofosu-Darkwah, & Adanu, 2018).

Self-efficacy figures prominently among several factors affecting behavior adoption (Bandura, 1977). Albert Bandura defined self-efficacy as perceived capability or belief that one can successfully complete a specific task or behavior (Bandura, 1977). Bandura differentiates between *outcome expectation*, i.e., the belief that a behavior will lead to a certain outcome, and *efficacy expectation*, i.e., the conviction that one can successfully execute the given behavior (Bandura, 1977) (Figure 1). For example, an efficacy expectation would be one’s confidence to successfully take intermittent preventive treatment of malaria in pregnancy (IPTp) three or more times during pregnancy, while an outcome expectation would be the belief that taking this treatment could prevent malaria in pregnancy. The environment must also enable the behavior, i.e., drugs to treat malaria in pregnancy must be available.

**Figure 1.**
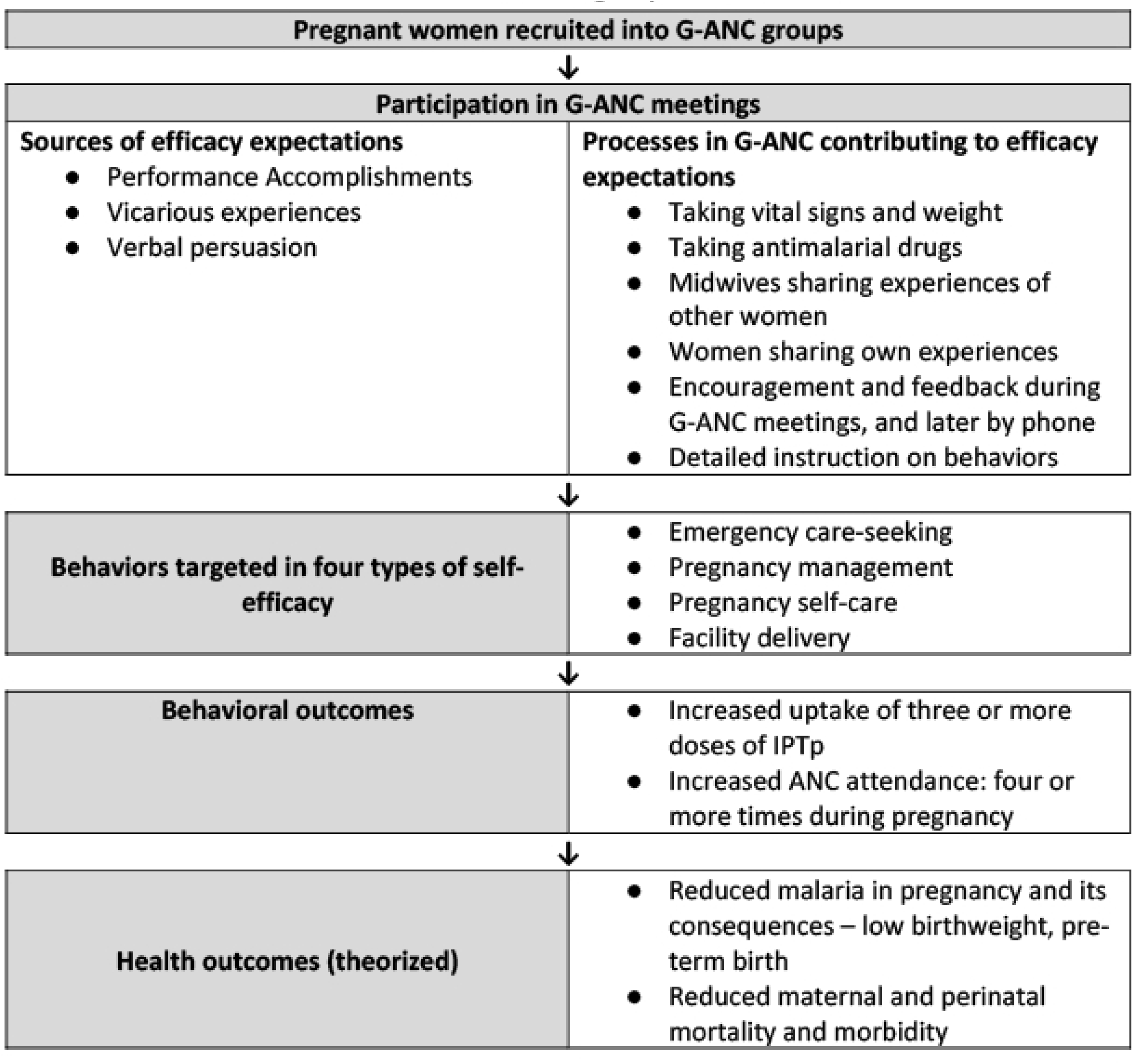
Processes, strategies and behavioral outcomes related to self-efficacy among pregnant women in group antenatal care.

Theoretically, G-ANC processes (e.g., taking and recording vital signs of other participants, learning from modeling experiences of others similar to oneself [Table 1 and Figure 1]) should strengthen participants’ self-efficacy by influencing Bandura’s four sources of self-efficacy expectations: 1) performance accomplishments, 2) vicarious experience , 3) verbal persuasion, and 4) emotional/physiological arousal (Bandura, 1977).

A nested qualitative study was conducted alongside a cluster randomized controlled trial assessing the impact of G-ANC on IPTp and ANC attendance in Atlantique Department, Benin (Gutman, 2024) to: 1) describe pregnant women’s experience of participation in G-ANC; 2) examine ways G-ANC might foster pregnant women’s self-efficacy to carry out recommended behaviors using Bandura’s four sources of efficacy expectations (12); and 3) identify ways G-ANC might foster changes in women’s propensity to adopt or maintain preventive and care-seeking behaviors.

## Methods

Quantitative and qualitative data collection took place from November 2, 2020 to December 15, 2022.

### Group Antenatal Care

From March 2021-September 2022, pregnant women attending individual ANC before 25 weeks’ gestation were invited to join a group of 8-15 women with similar due dates for monthly G-ANC meetings at their health facility instead of individual ANC (J. Sharma et al., 2018; Sheeder et al., 2012) (1, 2). Before the start of each meeting, women took each other’s blood pressure with a digital cuff and recorded this along with weight and temperature, sometimes assisted by a health aide. Every woman was individually examined by a provider in private, before or during the meeting.

After an opening ritual, the trained facilitator introduced the meeting topic from the G-ANC manual and led a participatory session where women interacted as a group and supported each other. The five-meeting series covered nutrition, malaria prevention, identification of danger signs in pregnancy, family planning, and preparation for birth. After covering the topic of the month, facilitators distributed sulfadoxine-pyrimethamine (SP) for IPTp under directly observed therapy. Each meeting ended with a closing song and dance.

### Household surveys

The parent study included baseline and endline household surveys in November-December 2020 and October-December 2022. Survey respondents were selected at random from a list of all women who had given birth in the preceding 12 months in one enumeration area per health facility catchment. The household survey covered questions for respondents in both arms on the parent study’s main outcomes of interest, IPTp uptake and ANC attendance, as well as questions adapted from the Malaria Behavior Survey Women’s Module (CCP, 2024) about ANC and perceived self-efficacy to attend ANC and take IPTp.

### Focus groups

From May-July 2022, two health facilities in each of Atlantique’s three Health Zones were purposively selected, for a total of six out of 20 intervention sites, to include one high (>100 clients/month) and one low (<100 clients/month) ANC client volume site per zone. Site selection also considered differing G-ANC enrollment and retention rates. A convenience sample of 140 women who attended G-ANC more than twice during their most recent pregnancy were invited to join focus groups. Ten focus groups were conducted by two trained interviewers in outdoor pavilions, open spaces, or health facility meeting rooms to understand women’s experience in their most recent pregnancy and with G-ANC, prevention of malaria in pregnancy, and family planning. Three intervention sites had multiple focus groups because of higher-than-expected participation.

### Data management and analysis

Focus groups were conducted in Fongbe, recorded, and transcribed into French in a template based on the focus group discussion guide. Results were coded and summarized in a Google Form by EG and PJW in consultation with FAEK and EFCD, following an adapted framework analysis process (Gale, Heath, Cameron, Rashid, & Redwood, 2013; Ward, Furber, Tierney, & Swallow, 2013). Deductive thematic analysis assessed women’s experiences and examined mechanisms through which participation in G-ANC promoted self-efficacy to adopt preventive and care-seeking behaviors based on Albert Bandura’s self-efficacy framework (Bandura, 1977). While pre-determined codes for role-playing, feedback, hands-on experience, and encouragement were applied in relation to Bandura’s four sources of self-efficacy (see Results), other aspects of G-ANC were identified as recurring themes.

Searches for self-efficacy statements included: 1) perceived ability to carry out behaviors, specifically looking for “can” and not “will” statements because confidence and capability are predictors of intention, not equivalent to it (Bandura, 1977); 2) reports or evidence of completion of tasks; and 3) signs of confidence and perceived capability.

Household survey data were collected in the CommCare platform, exported, and analyzed with SAS version 9.4 (SAS Institute Inc., Cary, NC, USA).

### Ethical considerations

The Institutional Review Board of the Centers for Disease Control and Prevention (ID 7254) and the Ethics Committee of the Benin Ministry of Health (ID 3/MS/DRFMT/CNERS/SA) approved the protocol prior to implementation of study activities. All data collectors completed the CITI online course on human subjects research. Community sensitization ensured that local leaders and community members were informed about the study. Health Zone officials and facility in-charges provided permission for study activities in each health center. Written informed consent was obtained from each person prior to their participation in G-ANC, surveys, or semi-structured interviews. Patients or the public were not involved in the design, conduct, reporting, or dissemination plans of this research.

## Results

Of 140 women invited to participate in focus group discussions, 129 participated. Nearly all focus group participants were married, averaged 26.7 years old, and had a mean of 2.8 children. Participants in most focus groups said they preferred G-ANC over individual ANC based on experience.

Women described receiving social support for their continued participation in G-ANC that helped them find solutions to such barriers to care as transportation costs, consultation fees, and hesitant spouses. They felt more confident advocating for themselves when faced with stressful situations (Table 2).

> “If you have a problem at home, you can tell [the midwife] and she’ll see what to advise you so that you can restore the peace at home. [After] the disagreements that existed at home, respect has settled into the home now; that too made me happy.”

Focus group participants reported that if a woman had financial difficulties that prevented her from travelling to the health center to attend a G-ANC meeting, or from paying the ANC fees, she could inform her peers, and they did what they could to help:

> “There are some people, because of the [costs of] travel [to the health center], they will say that the place is too far, they will say this and that. If you can’t find [money to pay for transport] and you call your second [the other woman in your pair], if she really has it [transportation money] or even if she doesn’t have it, she will try to help you find a solution…”

When a woman was absent, the provider and her peers contacted her, and tried to assist with the problems she was facing. This might include home visits by her peers, advice, provision of financial assistance, waiving of consultation fees by the midwife, or conflict mediation with her husband.

### Content domains of self-efficacy

We identified four content domains of self-efficacy that G-ANC processes targeted: 1) emergency care-seeking, 2) pregnancy management, 3) pregnancy self-care, and 4) facility delivery (Table 2). Higher self-efficacy for emergency care-seeking improved women’s autonomy to seek medical attention quickly, even if their husband’s support was needed to access care:

> “…it’s me who feels the pain who must say ‘I must go to the hospital’ before my husband tells me to go to the hospital, and ‘give me some money so that I can go to the hospital.’”

In the domain of pregnancy management, G-ANC increased participants’ confidence to monitor, interpret, and take appropriate action in response to pregnancy symptoms, and provided pregnant women assurance that they could handle pregnancy, childbirth, and newborn care:

> “Yes, I’m more confident that if I get pregnant, I could take care of the pregnancy right up to the time of delivery.”

G-ANC participants discussed improved self-efficacy to take IPTp, an aspect of pregnancy self-care. Many women reported that during previous pregnancies, they did not take prescribed medications and supplements. Women reported feeling more confident about taking iron supplements as well as IPTp, and making greater efforts to take medications while enrolled in G-ANC:

> “Yes, I take all [of the drugs offered at the health center]… At the beginning, I had difficulty taking the drugs, but as I started attending the meetings, the teachings, I started making the effort to take the drugs.”

In the fourth content domain, women described a greater ability to reach the health facility in time to give birth in a facility:

> “When the thing of the baby is going to start to sting [when contractions start], even if your husband doesn’t have any money, you’ll quickly get a motorcycle taxi to get to [the health center]… sometimes, because of money, things that shouldn’t happen do happen. So, we’ve been told that we have to buy a box and we’ll be contributing a little, a little money in there. After that, we have to quickly look for the motorcycle taxi that will take us [to the health center] because there are some people who it’s only when the baby thing starts to sting them that they start looking for a motorcycle and this ends up leading to other things [complications] and the child may no longer be found [the child may be stillborn], or the child’s mother may no longer be found [the mother may die].”

### Sources of efficacy expectations: Performance Accomplishments

G-ANC generated performance accomplishments when women experienced success at measuring and recording weight and vital signs, taking IPTp during meetings, and trying different behaviors at home that they learned in the group. For new behaviors performed outside of meetings, G-ANC participants reported back to the group on the recommended behaviors they had practiced on their own at subsequent meetings.

G-ANC participants reported pride in acquiring skills that made them feel more confident and valuable to the community. For example, women expressed self-satisfaction regarding how they had learned to take each other’s vital signs:

> “What I have gained in it, I myself know how to take the temperature of my neighbor, I myself know how to measure weight, how to take blood pressure, I know all this, whereas I didn’t know how to do anything before.”

### Sources of efficacy expectations: Vicarious experience

Vicarious experience during G-ANC came from watching health aides and other women in the group take one another’s vital signs, watching others take IPTp, role-playing, and sharing personal experiences. Women alluded to vicarious experience as one of the benefits of G-ANC, along with cohesion and solidarity within groups, and mutual aid between group members:

> “Before, when I came [to the health center], I felt alone, I sat alone and when my turn came, I would go to the midwife, and she would examine me. But now, the atmosphere is different, you feel like family, you have sisters.”

During time for sharing, women spoke of acquaintances who had poor pregnancy outcomes because they failed to address danger signs; participants offered advice from their previous pregnancies, and women who returned to the group after childbirth explained how lessons from G-ANC had helped them.

### Sources of efficacy expectations: Verbal persuasion

Because frequent ANC attendance throughout pregnancy is considered a key behavior to achieve proper pregnancy management, helping women overcome barriers to attendance is crucial.

Encouragement by peers to attend G-ANC meetings motivated participants to attend more ANC:

> “When you arrive at a [G-ANC] meeting, and you don’t see your second [the other woman in your pair] with whom you meet, when you start phoning them and you call them until… They always end up making themselves available, and we all come together.”

Participants explained that they also received advice from trusted providers who facilitated G-ANC:

> “When I took [IPTp] for the first time and I vomited; [the midwife] told me, if I have to take it another time, to not eat anything before taking it, and I followed that advice.”

Focus group participants consistently remarked on how much more time and care their providers took to give them detailed advice and explanations during G-ANC meetings, as compared to individual ANC.

### Sources of efficacy expectations: Emotional and physiological states

Focus groups did not describe stress or anxiety around taking IPTp and practicing other behaviors. They did however describe a positive emotional state induced by G-ANC that may have helped them to assimilate new information presented during meetings (Bandura, 1982; Totawar & Nambudiri, 2014).

### Survey Data on Self-Efficacy, Outcome Efficacy, and Comparison to Qualitative Findings

Across 20 intervention sites, 2,516 women attended at least one G-ANC meeting. The endline survey identified 140 of these participants out of a random sample of 1,280 households and found G-ANC participants were 1.9 times more likely than those who received only individual ANC to have attended four or more ANC visits (p=0.002) and 1.8 times more likely to have taken three or more IPTp doses (p=0.004).

Nearly all (96%; 134/140) of the women in the endline household survey who reported having participated in G-ANC said they would choose G-ANC again if they had another pregnancy. Women noted that they preferred G-ANC to individual ANC because they enjoyed being part of a group (74%), felt they received more comprehensive care (63%), and appreciated developing a relationship with one provider (49%).

More women who participated in G-ANC than those who had individual ANC reported that it was “somewhat true” or “mostly true” that they were given an opportunity to ask questions about family planning (p-value < 0.01) and had all their questions answered in a way they could understand (p-value = 0.02; Table 3). Despite focus group participants’ higher satisfaction with the education and advice from their providers compared with those who attended individual ANC, only 30.7% of endline survey respondents reported that it was “somewhat true” or “mostly true” that all their questions were answered in G-ANC, and only 28.7% said their questions were answered in a way they could understand. Focus group participants nevertheless described improvements in their understanding and comfort level with key behaviors. Some women reported that the drug given for IPTp was bitter, and pills had been difficult to swallow or caused undesirable effects. After confiding in their healthcare providers during G-ANC, they received coaching that helped minimize these effects and facilitated behavioral adoption.

Endline survey analysis found no significant difference between individual ANC and G-ANC participants when it came to health behaviors that they believed they had the self-efficacy to perform, such as successfully asking for IPTp (p-value 0.18) and taking IPTp at least three times during pregnancy (90 vs. 93.3%, p-value 0.29, Table 5). Belief in the outcome efficacy of taking IPTp was high whether or not women had participated in G-ANC (p-value 0.64, Table 5), as were other outcome efficacy beliefs. The only significant difference found was in recognizing the importance of taking IPTp while also sleeping under a bed net consistently (100% of G-ANC participants vs. 95.6% of individual ANC participants [p-value 0.01, Table 5]).

Self-efficacy beliefs about IPTp were higher than self-efficacy beliefs about accessing ANC in general, by 10 percentage points. This emerged in the household surveys when women were asked if they believed they could go for ANC as soon as they thought they might be pregnant (81.2%, 74.7-87.7) and if they believed they could convince their partners to accompany them to the health facility for ANC (76.1%, 70.6-82.9), as compared to results for their belief in the ability to take all three doses of IPTp (91.7%, 87.3-97.7) and in their ability to request IPTp at ANC (86.8%, 81.4-95.7).

## Discussion

Grenier et al. theorized that pride in new knowledge, skills, and the ability to act, along with increased social capital and support gained from G-ANC meetings, could lead to improved self-care and care-seeking practices (Grenier et al., 2022). This aligns with benefits of participating in G-ANC cited by participants in Benin, including group cohesion and solidarity, a positive emotional state while attending meetings, provider support, mutual aid between group members, and exposure to beneficial behaviors like taking IPTp. Opportunities in G-ANC to take blood pressure, observe other participants, and receive verbal persuasion may have provided sources of self-efficacy that empowered women to overcome barriers and perform recommended behaviors.

Similar to past studies from 16 low- and middle-income countries indicating G-ANC provides a better experience of care overall (Lazar, Boned-Rico, Olander, & McCourt, 2021; B. B. Sharma, Jones, Loxton, Booth, & Smith, 2018; J. Sharma et al., 2018), participants in Benin considered the care they received more comprehensive than in individual ANC. Women reported feeling that they developed both more detailed knowledge and greater confidence to perform health behaviors in four domains: emergency care-seeking; pregnancy management; pregnancy self-care; and facility delivery.

According to the parent study endline survey, over 98% of women in both study arms shared high outcome expectations that IPTp worked well to make sure one’s baby would be born healthy. Likewise, women’s self-efficacy to take IPTp at least three times during pregnancy was 90% even in the control arm, and focus groups did not express fear or anxiety as the main barriers to this behavior. Among pregnant women surveyed who participated in the G-ANC intervention, the parent study found a significant association with improved uptake of both ANC4 and taking three or more doses of IPTp. G-ANC in Benin was linked to a higher mean number of IPTp doses taken, compared with prior research in Nigeria and Kenya (Noguchi et al., 2020).

Endline survey results measured slightly higher outcome expectancy among G-ANC participants that taking IPTp in addition to sleeping under a mosquito net could prevent malaria. This finding points to knowledge gains; however, there was no significant difference in self-efficacy expectations among G-ANC and individual ANC participants when it came to convincing their partners to accompany them to ANC, asking for IPTp at the health facility, or taking at least three doses of IPTp while pregnant.

Evidence from the parent study and this sub-study suggest the factors that led G-ANC participants to take more IPTp during their pregnancies likely related to their accessing more ANC in general (Gutman, 2024) (J. Gutman et al., submitted manuscript). Social learning (i.e., learning behaviors through observation and modeling) entails a series of parallel processes (Bandura, 1977, 1982, 1998, 2001, 2004), and the positive emotional state that focus group participants reported during group meetings may have given women greater pleasure-based motivation to attend G-ANC (Totawar & Nambudiri, 2014). Persuasion by providers and peers to return for additional G-ANC meetings appears to be another component that led women to take more doses of IPTp.

While population-level coverage of G-ANC in Atlantique Department was low (14% in the intervention arm compared to a target of 50%), the endline household survey revealed that women who attended G-ANC had a more positive experience relative to their prior pregnancies, and 96% would prefer G-ANC over individual ANC in a future pregnancy. Studies in Nepal, Senegal, Bangladesh, and Rwanda had similar findings (McKinnon et al., 2020; Musabyimana et al., 2019; Sultana et al., 2019; Thapa et al., 2019).

Midwives who facilitated G-ANC meetings helped build participants’ self-efficacy during G-ANC meetings through hands-on practice, role playing, social support, and encouragement. They also induced a positive emotional state that may have helped reinforce new knowledge acquired through G-ANC meetings and encouraged more ANC contacts by G-ANC participants. In focus group discussions, women felt they had not only acquired knowledge, but also improved their capacity to overcome certain barriers to care. For instance, collective problem-solving and social learning similar to that described by Bandura (Bandura, 1982, 1998, 2001, 2002, 2004) helped women strengthen their ability to convince partners to let them attend regular ANC visits and seek care for pregnancy danger signs. G-ANC participants were able to find help if they needed it by confiding in providers and peers. Social support from G-ANC helped them to find ways of accessing care at the health facility. Participants described enhanced self-advocacy and exercising autonomy to overcome barriers to preventive care and care-seeking (e.g., saving their own money for facility-based childbirth). The knowledge and confidence gained were essential given the cultural context in which a woman often relies on her husband’s knowledge and opinions regarding health care. Still, participants remained less confident in their ability to convince partners to attend ANC with them than in their ability to perform other pregnancy-related behaviors that they could do on their own.

Social bonds formed between G-ANC participants and group facilitators emerged as a key benefit of G-ANC as a service delivery model. Consequently, G-ANC participants were more likely to seek providers’ help in navigating and managing their pregnancies. Focus group participants in Ghana reported feeling that G-ANC facilitators answered their questions completely (Lori et al., 2024); while some focus group participants in Benin highlighted getting more comprehensive answers to their questions during G-ANC meetings, this was not the case for respondents to the endline household survey for whom only one-third had all of their family planning questions answered. Participants in focus group discussions in Benin still said they learned and applied many new practices in G-ANC, which they had not known or done during past pregnancies.

### Study limitations

Interviews and focus groups were not designed specifically to investigate the effect of G-ANC on women’s self-efficacy. Self-efficacy questionnaires were not administered to all G-ANC participants prior to starting the intervention, so it is impossible to conclude through household survey results alone whether participants may have had higher self-efficacy prior to G-ANC participation or if they differed from non-participants in other qualitative ways. Moreover, the endline survey found evidence that most eligible women were not given the option to participate in G-ANC by their provider.

Bandura’s definition of self-efficacy focuses on word choice to distinguish between perceived capability and intention. Bandura emphasizes the difference between “will” and “can,” with “will” signaling intent and not self-efficacy, while “can” indicates perceived capability. The Fongbe verb *sixu* does not map directly onto the French verb *pouvoir* or the English verb *to be able*. Therefore, self-efficacy statements identified in the focus group transcripts may not accurately reflect Bandura’s English-language criteria for such statements.

## Conclusions

Prior to this study, G-ANC acceptability among pregnant women was unknown in the Beninese context. We found G-ANC to be favored by women in Atlantique Department who participated. We identified four domains of self-efficacy strengthened by G-ANC participation: emergency care-seeking; pregnancy management; self-care; and facility delivery. Although self-efficacy to take IPTp was already high at baseline and thus needed less reinforcement, processes inherent to G-ANC seem to have strengthened self-efficacy in other domains. G-ANC processes that empowered women to overcome barriers to ANC attendance and provided social support and encouragement to return to ANC (i.e., higher self-efficacy to seek care) may have facilitated their taking more doses of IPTp. While participants in G-ANC reported more opportunities to ask questions than those in individual ANC, future G-ANC training should emphasize that facilitators need to allow more time for questions in group and individual exam segments, answer in a way that women can understand, and focus on the parallel processes of G-ANC meetings that foster self-efficacy.

## Data Availability

The plan for data availability is currently under review by the institutions that participated in this study.

## Acknowledgements

The authors would like to acknowledge the health care workers, supervisors, and pregnant women who participated in this study, as well as 18 research assistants, the Benin Ministry of Health, Health Zone authorities (Abomey-Sô-ava, OKT, Allada-Toffo-Ze), and Shelby Cash of CDC PMI.

## References

Bandura, A. (1977). Self-efficacy: Toward a unifying theory of behavioral change. Psychological Review, 84(2), 191–215. 10.1037/0033-295X.84.2.191

Bandura, A. (1982). Self-efficacy mechanism in human agency. American Psychologist, 37(2), 122. 10.1037/0003-066X.37.2.122

Bandura, A. (1998). Health promotion from the perspective of social cognitive theory. Psychology & Health, 13(4), 623–649. 10.1080/08870449808407422

Bandura, A. (2001). Social Cognitive Theory: An Agentic Perspective. Annual Review of Psychology, 52(1), 1–26. 10.1146/annurev.psych.52.1.1

Bandura, A. (2002). Social Cognitive Theory in Cultural Context. Applied Psychology, 51(2), 269–290. 10.1111/1464-0597.00092

Bandura, A. (2004). Health Promotion by Social Cognitive Means. Health Education & Behavior, 31(2), 143–164. 10.1177/1090198104263660

CCP. (2024). Malaria Behavior Survey. Johns Hopkins Center for Communication Programs. Retrieved from https://malariabehaviorsurvey.org/.

Eluwa, G. I., Adebajo, S. B., Torpey, K., Shittu, O., Abdu-Aguye, S., Pearlman, D., . . . Chiegli, R. (2018). The effects of centering pregnancy on maternal and fetal outcomes in northern Nigeria; a prospective cohort analysis. BMC Pregnancy and Childbirth, 18(1), 158. 10.1186/s12884-018-1805-2

Gale, N. K., Heath, G., Cameron, E., Rashid, S., & Redwood, S. (2013). Using the framework method for the analysis of qualitative data in multi-disciplinary health research. BMC Medical Research Methodology, 13, 117. 10.1186/1471-2288-13-117

Grenier, L., Onguti, B., Whiting-Collins, L. J., Omanga, E., Suhowatsky, S., & Winch, P. J. (2022). Transforming women’s and providers’ experience of care for improved outcomes: A theory of change for group antenatal care in Kenya and Nigeria. PLoS One, 17(5), e0265174. 10.1371/journal.pone.0265174

Grenier, L., Suhowatsky, S., Kabue, M. M., Noguchi, L. M., Mohan, D., Karnad, S. R., . . . Smith, J. M. (2019). Impact of group antenatal care (G-ANC) versus individual antenatal care (ANC) on quality of care, ANC attendance and facility-based delivery: A pragmatic cluster-randomized controlled trial in Kenya and Nigeria. PLoS One, 14(10), e0222177. 10.1371/journal.pone.0222177

Gutman, J. R. (2025). Assessing the impact of group antenatal care on uptake of intermittent preventive treatment for malaria in pregnancy in Atlantique Department, Benin, 2021-2023: a cluster randomized controlled trial. Submitted.

Lazar, J., Boned-Rico, L., Olander, E. K., & McCourt, C. (2021). A systematic review of providers’ experiences of facilitating group antenatal care. Reproductive Health, 18(1), 180. 10.1186/s12978-021-01200-0

Lori, J. R., Chuey, M., Munro-Kramer, M. L., Ofosu-Darkwah, H., & Adanu, R. M. K. (2018). Increasing postpartum family planning uptake through group antenatal care: a longitudinal prospective cohort design. Reproductive Health, 15(1), 208. 10.1186/s12978-018-0644-y

Lori, J. R., Kukula, V. A., Liu, L., Apetorgbor, V. E. A., Ghosh, B., Awini, E., . . . Williams, J. (2024). Improving health literacy through group antenatal care: results from a cluster randomized controlled trial in Ghana. BMC Pregnancy and Childbirth, 24(1), 37. 10.1186/s12884-023-06224-x

McKinnon, B., Sall, M., Vandermorris, A., Traore, M., Lamesse-Diedhiou, F., McLaughlin, K., & Bassani, D. (2020). Feasibility and preliminary effectiveness of group antenatal care in Senegalese health posts: a pilot implementation trial. Health Policy and Planning, 35(5), 587–599. 10.1093/heapol/czz178

Musabyimana, A., Lundeen, T., Butrick, E., Sayinzoga, F., Rwabufigiri, B. N., Walker, D., & Musange, S. F. (2019). Before and after implementation of group antenatal care in Rwanda: a qualitative study of women’s experiences. Reproductive Health, 16(1), 90. 10.1186/s12978-019-0750-5

Noguchi, L., Grenier, L., Kabue, M., Ugwa, E., Oyetunji, J., Suhowatsky, S., . . . Adetiloye, O. (2020). Effect of group versus individual antenatal care on uptake of intermittent prophylactic treatment of malaria in pregnancy and related malaria outcomes in Nigeria and Kenya: analysis of data from a pragmatic cluster randomized trial. Malaria Journal, 19(1), 51. 10.1186/s12936-020-3099-x

Oka, M., Madeni, F., & Horiuchi, S. (2022). Effects of prenatal group program in rural Tanzania: A quasi-experimental study. Japan Journal of Nursing Science, 19(4), e12502. 10.1111/jjns.12502

Patil, C. L., Klima, C. S., Leshabari, S. C., Steffen, A. D., Pauls, H., McGown, M., & Norr, K. F. (2017). Randomized controlled pilot of a group antenatal care model and the sociodemographic factors associated with pregnancy-related empowerment in sub-Saharan Africa. BMC Pregnancy and Childbirth, 17(Suppl 2), 336. 10.1186/s12884-017-1493-3

Sharma, B. B., Jones, L., Loxton, D. J., Booth, D., & Smith, R. (2018). Systematic review of community participation interventions to improve maternal health outcomes in rural South Asia. BMC Pregnancy and Childbirth, 18(1), 327. 10.1186/s12884-018-1964-1

Sharma, J., O’Connor, M., & Rima Jolivet, R. (2018). Group antenatal care models in low- and middle-income countries: a systematic evidence synthesis. Reproductive Health, 15(1), 38. 10.1186/s12978-018-0476-9

Sheeder, J., Weber Yorga, K., & Kabir-Greher, K. (2012). A review of prenatal group care literature: the need for a structured theoretical framework and systematic evaluation. Maternal and Child Health Journal, 16(1), 177–187. 10.1007/s10995-010-0709-1

Somji, A., Ramsey, K., Dryer, S., Makokha, F., Ambasa, C., Aryeh, B., . . . Rashid, S. (2022). “Taking care of your pregnancy”: a mixed-methods study of group antenatal care in Kakamega County, Kenya. BMC Health Services Research, 22(1), 969. 10.1186/s12913-022-08200-1

Sultana, M., Ali, N., Akram, R., Jahir, T., Mahumud, R. A., Sarker, A. R., & Islam, Z. (2019). Group prenatal care experiences among pregnant women in a Bangladeshi community. PLoS One, 14(6), e0218169. 10.1371/journal.pone.0218169

Thapa, P., Bangura, A. H., Nirola, I., Citrin, D., Belbase, B., Bogati, B., . . . Maru, S. (2019). The power of peers: an effectiveness evaluation of a cluster-controlled trial of group antenatal care in rural Nepal. Reproductive Health, 16(1), 150. 10.1186/s12978-019-0820-8

Totawar, A. K., & Nambudiri, R. (2014). Mood and self-efficacy: The moderation of hedonic and utilitarian motivation. Human Resource Development Review, 13(3), 314–335. 10.1177/1534484313492330

Ward, D. J., Furber, C., Tierney, S., & Swallow, V. (2013). Using Framework Analysis in nursing research: a worked example. Journal of Advanced Nursing, 69(11), 2423–2431. 10.1111/jan.12127

